# Comparison of VATS Pleurectomy/Decortication Surgery plus Hyperthermic Intrathoracic Chemotherapy with VATS talc pleurodesis for the treatment of Malignant Pleural Mesothelioma: a randomized pilot study

**DOI:** 10.1101/2021.11.27.21265291

**Authors:** Marcello Migliore, Maria Fiore, Rosario Tumino, Tommaso Filippini, Marco Nardini, Corrado Spatola, Sergio Castorina, Paolo Vigneri, Ines Monte, Riccardo Polosa, Margherita Ferrante, the University of Catania Study Group on Malignant Pleural Mesothelioma

## Abstract

**Introduction:** No previous study provides compelling evidence to convince surgeons to opt for one procedure over another for the treatment of Malignant Pleural Mesothelioma (MPM). Hyperthermic intrathoracic chemotherapy (HITHOC) adjunct to surgery for MPM has no definite role. The primary objective of this randomized pilot trial was to evaluate the feasibility for future large studies.

**Method:** The study design was a prospective randomized three-centric pilot trial (ISRCTN12709516). We recruited patients diagnosed with MPM and prospectively assigned them to two groups: Group A Video Assisted Thoracic Surgery (VATS) talc pleurodesis or Group B VATS P/D plus HITHOC. The main outcome measures were description of study feasibility. We collected socio-demographic and clinical patient information. Data of Kaplan-Meier survival analysis and Cox regression analysis are presented.

**Results:** From November-2011 to July-2017 24 males and 3 females, with a median age of 68-years were enrolled (recruitment rate 5 patients/year). Preoperative stage was I-II, and 18 had epithelioid type. 14 patients were in the Group A. Operative mortality was 0. Follow-up ranged from 6 to 80 months. The median overall survival time started to diverge at 20 months. In Group A, it was 19 months (95% CI:12-25) and in Group B, **it** was 28 months (95% CI:0-56). Survival rate for the epithelioid type was 15 months (0-34) in groups A. and 45 months (0-107) in the Group B with an HR 0.77 (95% CI:0.28-2.2).

**Conclusion:** These findings suggest that VATS P/D plus HITHOC may improve survival time in MPM patients undergoing surgical treatment and support the need for a larger multicenter randomized clinical trial.

**Strengths and limitations of this study:**

- this pilot study represents an important step forward of the treatment of malignant pleural mesothelioma
- The study demonstrates the feasibility for a multicenter randomized trial to compare VATS P/D plus HITHOC with VATS talc pleurodesis in the management of MPM
- Although survival for the epitheliod type is 45 months, the inclusion of small number of patients is a limitation.
- Although neoadjuvant, adjuvant chemo-radio or immunotherapy have been administered to the patients, we have no specific information about doses for every single patient.
- A potential source of bias could be that the house staff of the Centre A has not been formally educated about the study while in the Centre B house staff was informed about the pilot trial.

## Introduction

Malignant Pleural Mesothelioma (MPM) is a rare but very aggressive cancer arising from the pleura often diagnosed in a locally advanced stage. Its worldwide incidence has been increasing. Total incidence is highest in the USA and UK although per capita, Australia and Italy also rank highly.^1^ Despite the main risk factor for mesothelioma is occupational exposure to asbestos, environmental exposure to these fibers also plays an important role and both contribute to the formation of clusters.^2,3^

The prognosis of MPM is poor and the mean survival period of the patients is 9-12 months^1^. Analysis of Mesothelioma mortality recorded in the WHO mortality database between 1994 and 2008 yielded an age-adjusted mortality rate of 4.9 per million, a mean age at death of 70 years and male to female ratio of 3.6:1.^4^

Treatments options are often limited to palliative care and only few have intention to cure. The type of treatment depends on center or surgeon experience, cancer staging and patient’s individualities such as age and performance status. There is abundant evidence to affirm that pleuropneumonectomy is nowadays rarely performed because the reported higher mortality^5^ and because similar or even better long-term results which can be obtained with less invasive surgeries such as pleurectomy/decortication (P/D) or talc pleurodesis (TP) alone.^6,7^ Pleurectomy/decortication is a lung sparing resection which permits the macroscopic removal of the tumor, and has a low surgical mortality rate with a significant risk of local recurrence. Furthermore, the mean survival for patients treated with TP alone is 14 months, and P/D, which shows a mortality of 1.8%, has a reported survival of 17 months.^6,7^ Hyperthermic Intraoperative Thoracic Chemotherapy (HITHOC) is a type of adjuvant local treatment performed in the operating room immediately after surgery for MPM, and many experiences and studies have been reported to date a median survival rate ranging from 20 to 35 months.^8^ Nonetheless, there is no gold standard treatment for MPM, and therefore the challenge for the future is to develop strategies that will bring good quality of life, longer disease-free interval and prolonged overall survival.

## Objective

The primary objective of this randomized pilot trial was to evaluate the feasibility for a larger randomized study comparing surgical treatments for MPM.

### Primary outcome measures

1. Description of study feasibility (recruitment and retention rates)

### Secondary outcome measures

1. Survival rates of participants at 1, 2, 3 years were recorded through personal interview or phone calls during the trial period.
2. Length of hospital stay was assessed using case report forms throughout the period of hospitalization.
3. Perioperative and late complications from the procedure were assessed at baseline, 3, 6, and 12 months.
4. Estimate of the sample size required for final trial
5. Estimate of the number of recruiting centers required for final trial

## Methods

### Study design

This study is designed as prospective parallel-groups randomized three-centric pilot trial (ISRCTN12709516), with allocation ratio 1:1.

After the first year we changed randomization strategy (see Randomization chapter).

### Participants, setting

The study participants were patients diagnosed with Malignant Pleural Mesothelioma. Five Sicilian Thoracic centers have been contacted and three centers agreed to participate to the pilot trial. This pilot trial was conducted at the Thoracic surgery at Morgagni Hospital (Center A1) and University of Palermo Hospital (Center A2), at the University of Catania Hospital (Centre B). All patients and survival have been validated by the COR Sicily (Regional Operative Centre for the Italian National Mesothelioma Registry) which is a dedicated Institution of the Italian Government.^9^

#### Patient and public involvement

Patients or the public were not involved in the design, or conduct, or reporting, or dissemination plans of our research

### Inclusion criteria

1. Patients who gave written consent to participate.
2. Patients with MPM with or without pleural effusion, with a performance status equal or below 2.
3. Patients who agreed to undergo VATS talc pleurodesis

### Exclusion criteria

1. Patients unfit for a VATS procedure.
2. Clinical evidence of disease progression since diagnosis to advanced stage (stage III-IV).

Between November 2011 to July 2017 all physicians and surgeons of the participant centers identified patients with MPM that met the inclusion criteria. Patients were then approached by the operating surgeon who explained the operative procedures and the study. Then the informed written consent was obtained.

### Interventions

Participants received either VATS Pleurectomy/Decortication plus HITHOC or VATS talc pleurodesis, according to which participating Centre they attended. This was determined by the existing expertise at each of those participating centers. All patients who were admitted to Centers A1 and A2 underwent VATS talc pleurodesis, while all patients admitted to Centre B underwent VATS P/D plus HITHOC. Neoadjuvant, adjuvant chemo-radio or immunotherapy was administered.

#### The VATS talc pleurodesis arm (Group A)

Group A participants were admitted to the Morgagni Institute (Centre A) and underwent uniportal VATS talc pleurodesis^10^ drainage of the pleural effusion, biopsy and frozen section. When the results of the frozen section analysis demonstrated the presence of a thoracic cancer, primary or secondary, patients received the VATS talc pleurodesis to avoid the accumulation of excess fluid, talc being the most effective sclerosant agent for treatment of malignant pleural effusion. Under direct vision, talc was inserted onto the entire lung surface. A drain was kept on suction until drainage stopped in order to promote adhesions between the lung and chest wall.

#### The VATS-PD and HITHOC arm (Group B)

Group B participants were admitted to the University of Catania (Centre B) and underwent VATS P/D plus HITHOC. A mini-thoracotomy of 10-12 cm was performed and a 10 mm 0° optic was used to assist the surgeon. The debulking procedure is less invasive than a pleuropneumonectomy and includes pleurectomy (removal of parietal pleura), decortication (removal of visceral pleura) and removal of all tumour able to be seen by the human eye.^11-13^ HITHOC is a concentrated dose of chemotherapy with cisplatin diluted in a normal saline solution of 2-3 litres, warmed at 41°C, and infused and circulated in the chest for 60 minutes. When present, the pleural effusion was aspirated at the beginning of the operation to drain excess fluid from the chest. All randomized patients had 100 mL of pleural effusion taken for cytological analysis at the beginning of the operation, at the end of the operation and at the end of the HITHOC. *Figure 1* demonstrates the three steps of the VAS-PD HITHOC method.

**Fig 1.**
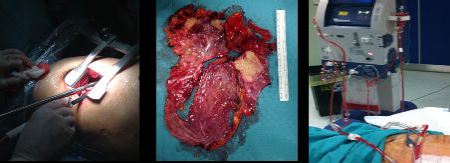
VATS P/D and HITHOC. a) mini-thoracotomy; b) pleurectomy; c) HITHOC

All patients have been followed throughout the study using personal interview and record card including assessments and measurements to address each pilot trial aim. Patients were instructed to attend the outpatient the clinic at 3, 6, 12 months and when necessary. No changes to pilot trial assessments or measurements after the pilot trial commenced have been done.

Feasibility for a larger randomized study was the main outcome of our pilot study. Priori criteria were the following: (a) the proportion of the invited centres accepting to participate would be 50% or greater; (b) the number of the patients recruited for each centre would be minimum of three or greater per year.

As this is a feasibility study a formal sample size calculation is not required, but we estimated the number of participants required by Julius’ criteria.^14^

### Randomization

Initially (November 2011), the study randomization was performed in-house using the envelopes method. During the first full year (2012) three patients were enrolled. Failure to recruit sufficient numbers became evident and we were not able to complete the study. We therefore contacted thoracic units in Sicily, and in January 2014 two Units joined the study. We then modified the randomization method according to the experience of each Centre. All patients with mesothelioma seen at the Morgagni Hospital and University Hospital of Palermo (Group A) were treated with uniportal VATS talc pleurodesis, whilst those undergoing surgery at the University Hospital of Catania (Group B) were treated with VATS Pleurectomy/Decortication plus HITHOC. The enrolment was closed July 31, 2017 and patients were followed-up until December 31, 2020. In both centers the eligible patients were identified by the oncologists and surgeons responsible of their clinical assessment.

### Statistical analysis

This is a pilot trial with the sample size based on feasibility. Given the lack of previous evidence of talc pleurodesis efficacy in terms of risk reduction, we decided to do a pilot study in small groups of 12 patients in each arm following the criteria of Julius and CONSORT extension to randomized pilot studies.^14,15^ We addressed each pilot objective using quantitative method. Continuous and categorical variables were expressed as median (interquartile range-IQR) and relative frequencies (%), respectively. The probability of survival over time was estimated using the Kaplan–Meier method and expressed as median and 95% confidence interval (CI). We calculated Hazard Ratios (HR) and 95% CI, with the Group A (VATS talc pleurodesis) as the reference, using a Cox proportional hazard regression model, adjusting for age, sex, histological subtype, asbestos exposure, performance status (1, 2), TNM, and STAGE (1a, 1b). We also carried out stratified analysis by smoke history as potential effect modifier. All analyses were on an intention-to-treat basis. Statistical analysis was performed using statistical software SPSS for Windows (Statistical Package for the Social Science, version 21.0; SPSS Inc., Chicago, IL, USA).

## Results

For each group, the numbers of participants who were assessed for eligibility, randomly assigned, received intended treatment, and assessed for each objective were reported in *Figure 2*.

**Fig 2.**
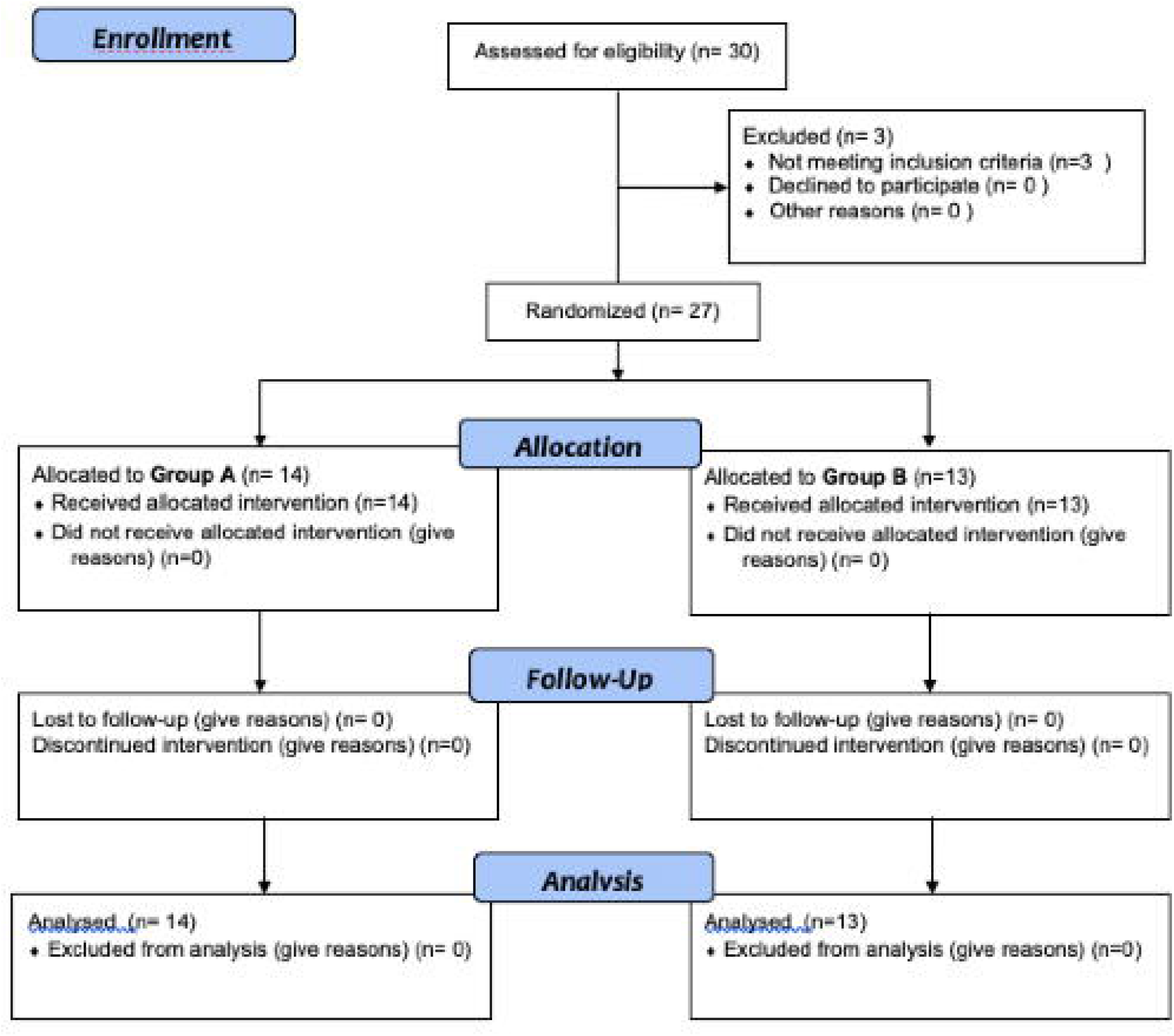
Patients flow from enrollment to analysis.

Patient enrolment started in November 2011 and was completed in July 31, 2017. Three-centers out of five agreed to participate (more than the proportion established as priori criterion above 50%) and five patients/year have been recruited. No patients have been lost during follow-up. The University Hospital of Palermo contributed with 3 patients who did not meet the inclusion criteria and have been excluded (Figure 2).

### Patients’ demographics

A total of 27 patients were included in the study: 24 were male and 3 were females (2 females in Group A and 1 in Group B). Median age of the combined groups was 68 years (IQR: 59-74). Socio-demographic and clinical patient’s characteristics and staging (according to TNM Classification of Malignat Tumours, 7^th^ Edition)^16^, by Groups are shown in *Table 1*. Eleven out 27 patients (40.7%) reported asbestos exposure. Fourteen patients (51.8%) were smokers. Seven patients of the Group B underwent neo-adjuvant treatment. All patients had adjuvant treatment.

**Table 1:**
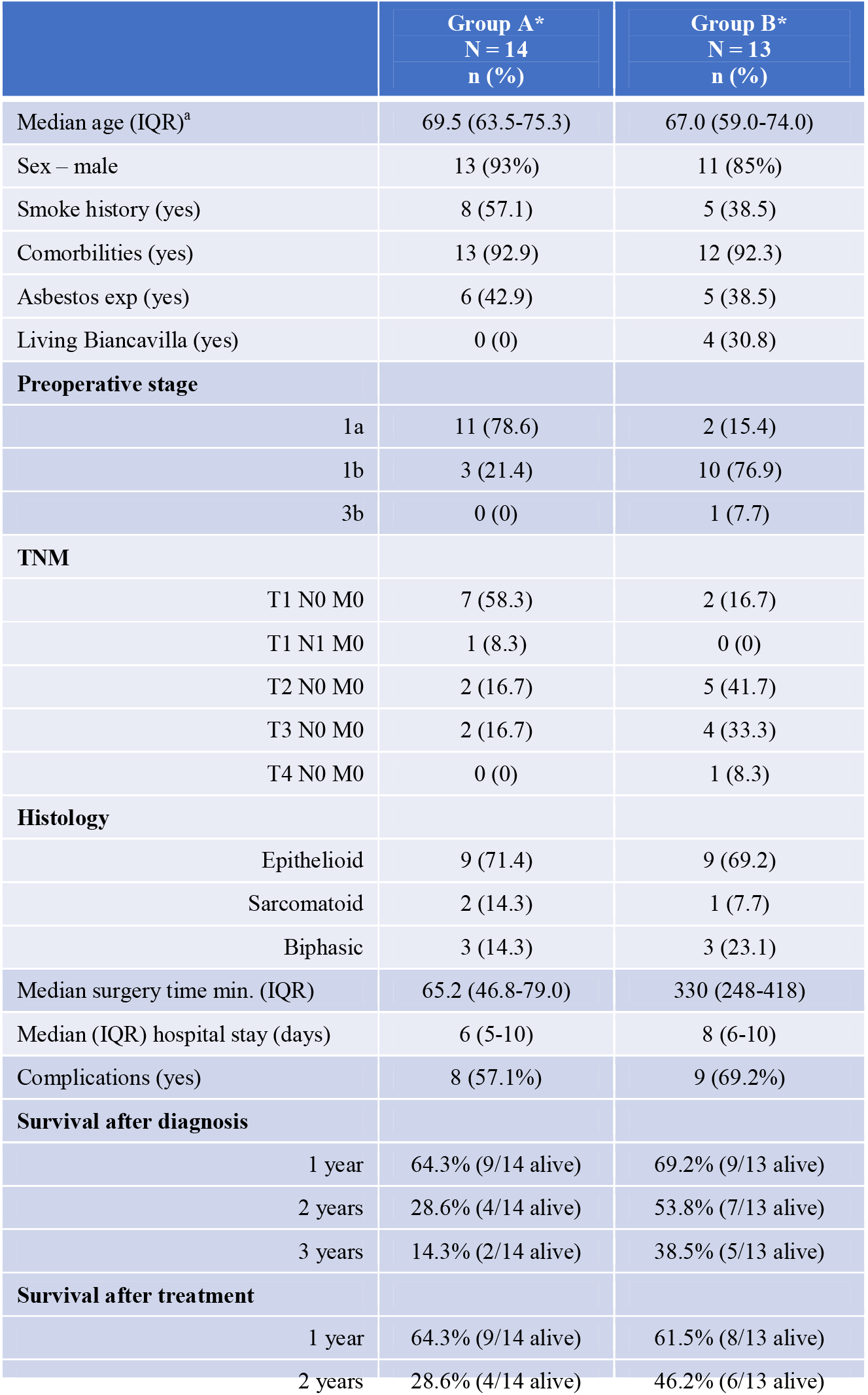
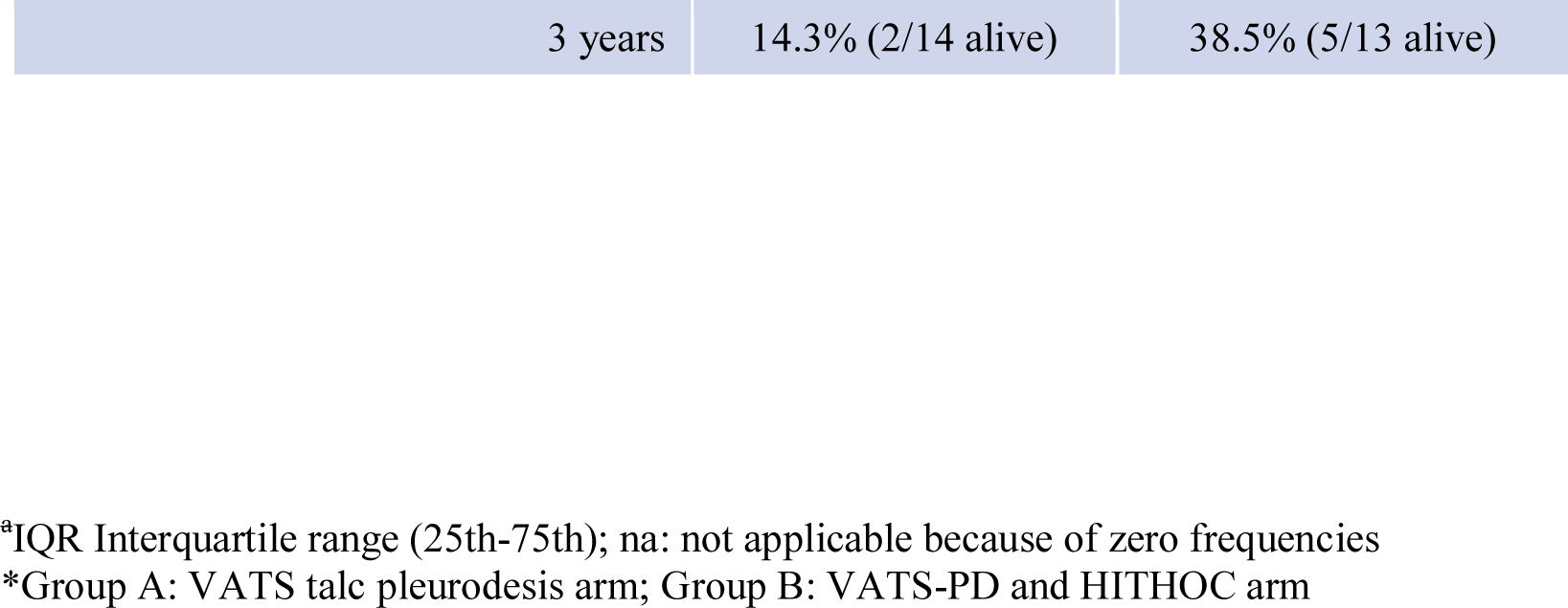
Socio-demographic and clinical patient’s characteristics, by groups

#### Group A

(VATS talc pleurodesis alone) consisted of 14 patients (1 female, 13 males) with a median age of 70 years (IQR 64-75). In this cohort there were 6 patients (42.8%) who reported asbestos exposure, and 9 (57.1%) were confirmed as smokers.

The symptoms at diagnosis they reported were: cough, dyspnea, chest pain, odynophagia, asthenia, shoulder pain, dysphonia, and weight loss; one patient refers no symptoms.

Computed Tomography (CT) and Positron Emission Tomography (PET) scans were carried out in all patients after surgery.

All patients of Group A underwent surgery for recurrent undiagnosed pleural effusion. Final diagnosis of MPM confirmed 9 patients with Epithelioid MPM, 3 with biphasic and 2 with sarcomatoid. According to preoperative pTNM staging, all patients of Group A were in stage IA or IB.

#### Group B

(VATS P/D plus HITHOC) consisted of 13 patients (2 females, 11 males), with a median age of 67 years (IQR 59-74). Five patients (38.5%) reported asbestos exposure, and 4 patients (30.7%) were from Biancavilla; five patients (35.5%) were confirmed as smokers. The symptoms reported were: cough, dyspnea, chest pain, asthenia, weight loss; one patient refers no symptoms.

CT and PET scans were carried out in all patients before surgery. All patients at the time of surgical treatment had a confirmed diagnosis of MPM - obtained in four patients via thoracentesis (cytological diagnosis), in two patients via CT-guided biopsy, in 7 patients after VATS-biopsy. Epithelioid MPM was diagnosed in 9 patients, biphasic in three and Sarcomatoid in one patient. The median time between diagnosis and surgery was 2 (IQR 1-6) months.

According to pTNM staging, all patients in Group B were in stage IA or IB, except one patient in stage IIIB (as assessed post-operation). Several wedge resections were performed in three patients, diaphragmatic reconstruction with a patch in two, and rib resection in one.

### Outcomes of the operations

Median surgery time for Group A was 65.2 min (IQR 46.8-79.0). Median operative time, from the time of skin incision to the end of perfusion for Group B, was 316 min (IQR 224-425 min). The median hospital stay was 6 (IQR 5-10) and 8 (IQR 6-10) days for Group A and Group B, respectively. There was no incidence of intra-operative complications in any of the 27 patients. Post-operative complication rate was 8 (51.7%) and 7 (69.2 %) in Group A and B, respectively (*Table 2*). In five patients of the group B the mini-thoracotomy was enlarged due to tight adhesions.

**Table 2.** Distribution of post-operative complications, by groups.

| **Group A\***<br>(N=14) | n (%) | **Group B\***<br>(N=13) | n (%) |
| --- | --- | --- | --- |
| Incomplete pulmonary expansion | 4 (28.6) | Anemia | 2 (15.4) |
| Melaena | 1 (7.1) | Tingle in the lips and nausea | 1 (7.7) |
| Hemothorax | 1 (7.1) | Postural and stay tremor | 1 (7.7) |
| Pleural effusion | 2 (14.3) | Tachyarrhythmia | 1 (7.7) |
| - | - | Syncope | 1 (7.7) |
| - | - | Persistent Air-leak | 1 (7.7) |

\*Group A: VATS talc pleurodesis arm; Group B: VATS-PD and HITHOC arm

Four patients (28.6%) of Group A presented with incomplete pulmonary expansion after uniportal VATS talc pleurodesis. No patients required further surgery to address post-operative complications. There was no hospital mortality for patients in either group. No mortality was documented at 30 and 60 days in either group. One patient, who is still alive at the time of writing this article, underwent two further operations for tumor recurrence.

### Follow-up

For Group A, with a median follow-up time of 19 months (IQR 9-32) from time of diagnosis, one out of 14 patients (7.1%) was still alive at the end of follow-up. For Group B, with a median follow-up time of 28 months (IQR 11-56) from time of diagnosis, four out of 13 patients (30.8%) were still alive.

### Survival rates of participants at 1, 2, 3 years

A complete overview of the follow-up after surgery for both groups at 3, 6, 12, 24, 36 months and dates about survival is shown in the *Table 3*.

**Table 3:** Survivors during follow-up (months), by groups.

| **Group** | **3 m** | **6 m** | **12 m** | **24 m** | **36 m** |
| --- | --- | --- | --- | --- | --- |
|  | **n(%)** | **n(%)** | **n(%)** | **n(%)** | **n(%)** |
| A | 14 | 12 | 9 | 3 | 1 |
| (14 patients) | 100% | 85,7% | 64% | 21% | 7.1% |
| B | 13 | 11 | 9 | 7 | 4 |
| (13 patients) | 100% | 84,6% | 69% | 54% | 30.7% |

\*Group A: VATS talc pleurodesis arm; Group B: VATS-PD and HITHOC arm

For the Group A survival after diagnosis and treatment was coincident while for the Group B survival after treatment did not correspond at the survival after diagnosis as treatment was performed from 0.5 to 6 months after diagnosis.

The most important difference in terms of survival from time diagnosis, even if imprecise, was detected after 20 months when the median overall survival started to diverge. The median overall survival time after diagnosis for Group A was 19 months (95% CI: 12-25) versus 28 months (95% CI: 0-56) of Group B; whereas median overall survival after treatment for Group A was 19 months (95% CI: 12-25) versus 23 months (95% CI: 0-50) of Group B (Figures 3a and 3b). The HR for overall survival, at time diagnosis, between groups A and B was 0.52 (95% CI: 0.22-1.23), and after adjustment for age, sex, histological subtype TNM (one patient stage T1N1M0 of group A and one patient T4N0M0 of the group B have not been included in the analysis because they were present only in one group), STAGE (1a, 1b; one patient staged 3b has not been included in the analysis because there was no corresponding patient in the other group) and asbestos exposure HR was 0.15 (95% CI: 0.02-0.91).

**Fig. 3a and 3b:**
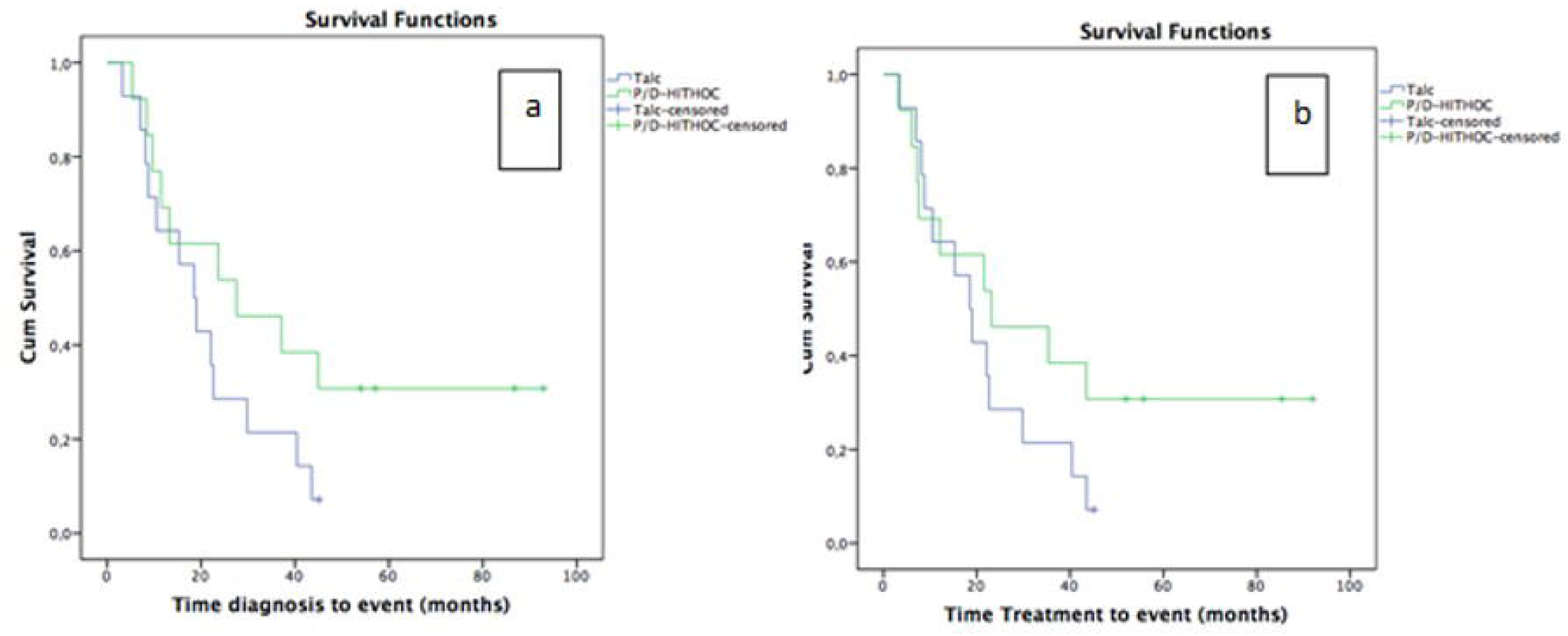
Kaplan-Meier curves comparing VATS talc pleurodesis (Group A) and VATS P/D-HITHOC (Group B) at time diagnosis and at time treatment.

Median overall survival time from diagnosis for non-smokers and smoker patients was 19 months for (95% CI: 1.6-36) and 15 months (95% CI: 4-26) for Group A versus 28 months (95% CI: 9-46) and 11 months (95% CI: 7-15) for group B, respectively (Figure 3a and 3b). Median overall survival time from treatment for non-smoker and smoker patients was 19 months (95% CI: 1.6-36) and 15 months (95%: CI 4-26) for Group A while it was 23 months (95% CI: 4-42) for non-smokers and 8 months (95% CI: 4-11) for smokers in Group B. In non-smoker patients, the risk was reported to be 65% lower (HR= 0.35, 95% CI 0.09-1.33).

Patients with epithelioid MPM histology showed a median survival of 15 months (IQR 0-34) in Group A and 45 months (IQR 0-107) in Group B. The presence of epithelioid MPM post diagnosis (HR=0.87, 95% CI 0.32-2.4) and after treatment (HR=0.77, 95% CI: 0.28-2.2), showed a risk of death lower than patients with biphasic histology (Figures 4a, 4b); whereas patients with sarcomatoid MPM histology both at the time of diagnosis (HR=1.84, 95% CI: 0.44-7.70) and after treatment (HR=1.62, 95% CI: 0.39-6.80) showed a risk of death higher than patients with a biphasic histology (*Table 4*).

**Table 4.** Median survival months (95% CI) and hazard ratio (HR) according to final histology, by groups.

| Histology | n | Time diagnosis |  | Time treatment |  | HR (95% CI)* |  |
| --- | --- | --- | --- | --- | --- | --- | --- |
|  |  | Group A** | Group B** | Group A** | Group B** | Time treatment | Time diagnosis |
| Biphasic | 6 | 30 (17-42) | 28 (1.8-53) | 30 (17-42) | 22 (0-46) | *Ref.* | *Ref.* |
| *Epithelioid* | 18 | 15 (0-34) | 45 (0-107) | 15 (0-34) | 43 (0-103) | 0.77 (0.28-2.2) | 0.87 (0.32-2.4) |
| *Sarcomatoid* | 3 | 11 (na) | 10 (na) | 11 (na) | 8 (na) | 1.62 (0.39-6.8) | 1.84 (0.44-7.7) |
| *Epitelioid\*\*\** |  | - | - | - | - | 0.80 (0.29-2.2) | 0.87 (0.31-2.4) |

\*95% Confidence interval
*na: not applicable because low frequencies*
\*\*Group A: VATS talc pleurodesis arm; Group B: VATS-PD and HITHOC arm
\*\*\* *excluding the sarcomatoid*

**Fig. 4a and 4b:**
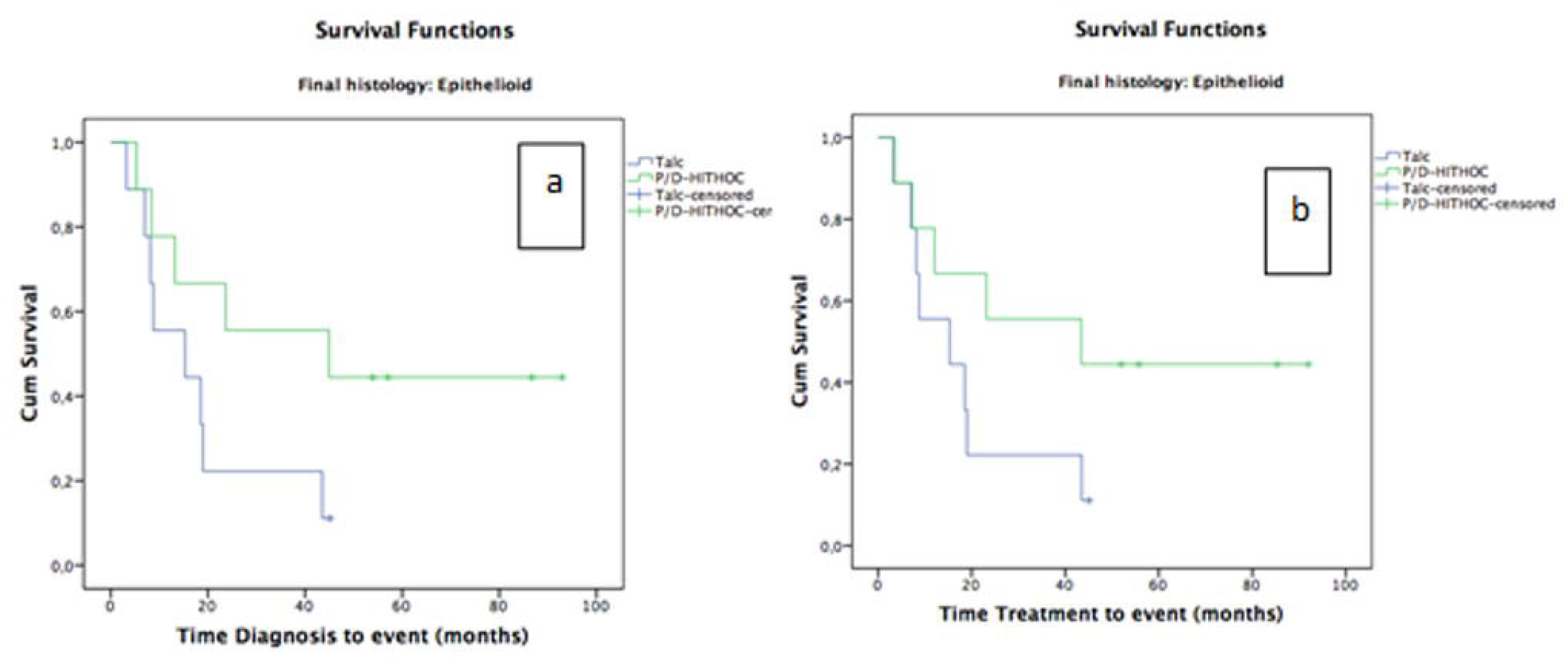
Kaplan-Meier curves comparing VATS talc pleurodesis (Group A) and VATS P/D-HITHOC (Group B) and at time diagnosis and time treatment for epithelioid mesothelioma.

### Sample size calculation for future trial

Considering the HR for overall survival found in this pilot trial of 0.52, the needed samples size for a future trial, based on alfa significance level of 0.05 and 80% of power^17^, should be of 145 patients with approximately 73 events/deaths.

## Discussion

After years of skepticism regarding surgical treatment of patient with mesothelioma, a more positive era began in the late 1980s and alternative surgical treatment protocols started to be trialed with the aim at improving surgical outcomes for this aggressive cancer. Unfortunately, all studies to date confirmed that the role of surgery remains very uncertain due, in part, to the lack of large prospective randomized trials, which might confirm its role.^18-20^

In truth, few randomized controlled trials comparing procedures for MPM, ranging from radical (EPP) to less radical VATS P/D or VATS talc pleurodesis have been done, and none of them represents the gold standard for the treatment of MPM.^5,6^

The questions on “why surgery does not work in Malignant Pleural Mesothelioma” (which was posed in an editorial of few years ago) remains relevant,^21^ and from reading the literature it is evident that extended operations for MPM such as pleuropneumonectomy should definitively be abandoned because long term survival is not enhanced, and in-hospital mortality is unacceptable when compared with those results obtained with less invasive approaches.^22^

Thoracic surgeons are aware that in mesothelioma even the most aggressive surgery treatment cannot guarantee removal of all residual cancer cells. The main objective of the HITHOC treatment has therefore been to completely eradicate any remaining cancer cells.^8^

A large study on VATS P/D plus HITHOC was carried out at Brigham and Women’s Hospital, Harvard Medical School, Boston, in 2013.^23^ It enrolled 103 patients with a diagnosis of MPM who were treated with VATS P/D via a thoracotomy: 72 patients received HITHOC (Group A) and 31 patients received no HITHOC (Group B). Group A showed an overall survival time of 35.3 months compared with 22.8 months for Group B. Although these results have not been confirmed by other centres, many groups have shown an important increase in survival with a good quality of life.^24-27^

Our study differs from the Harvard study and many others, as all our patients underwent VATS via a mini-thoracotomy of 10-12 cm instead of a large thoracotomy with rib resection. With this minimally invasive strategy, a low mortality has been confirmed along with a low complication rate, low general toxicity and a total preservation of respiratory function. Moreover, another reported advantage is that this minimally invasive technique could stimulate a positive immunological patient reaction.^28^

It is important to note that in both Group A and Group B of our study, the difference in the amount of time spent in surgery did not affect the duration of hospital stay or postoperative complications, demonstrating the safety of the combination of VATS P/D and HITHOC.

We calculated survival time after diagnosis and after treatment, and the overall survival time increased in favour of VATS P/D plus HITHOC with a median survival of 28 months and with 30% of patients still alive at the end of follow-up on December 31, 2020. This information is important as it reinforces that overall survival could be further prolonged, and also justifies the undertaking of a larger randomized-controlled trial.

It is also noteworthy that in Group B, the median survival time in the 9 patients with epithelioid MPM undergoing VATS P/D plus HITHOC, was 45 months, versus 15 months in the 9 patients with epithelioid MPM undergoing VATS talc pleurodesis. It is certainly not new that patients with epithelioid MPM survive longer than those with other types of MPM, but the long-term results from our study are encouraging for the future treatment of this aggressive cancer.

In the future study we should pay more attention to Biancavilla, town located in a volcanic area of eastern Sicily of special interest because of a cluster of mesothelioma cases,^3^ where our results suggest a longer survival for these patients. This could be due to the type of treatment (VATS P/D plus HITHOC), to the smoking habits, but also to a different carcinogenic effect of the fibre found in Biancavilla (Fluoroedenite).

**Limitations of our study** include the small number of patients. To overcome this limitation, the participation in the future study will also be offered both to national and international specialist centers. In our study, approximately more than half of the eligible centers agreed to the study. This information is important for planning the future trial and because the involvement of other centers would be helpful not only to increase the sample size but because this methodology could also be applied in research settings other than the future definitive RCT. Although neoadjuvant, adjuvant chemo-radio or immunotherapy have been administered to the patients, we have no specific information about doses for every single patient. Another limitation is that we did not know how long the patients had quit smoking, so we have to add to the questionnaire more specific questions regarding smoking habits. A potential source of bias could be that the house staff of the Centre A has not been formally educated about the study while in the Centre B house staff was informed about the pilot trial. This difference could be cause of better care in the Centre B. To avoid remaining uncertainty about feasibility, the described potential bias will be resolved informing the staff about the project.

In conclusion, this pilot study represents an important step forward demonstrating the feasibility for a multicenter randomized trial to compare VATS P/D plus HITHOC with VATS talc pleurodesis in the management of MPM, and to estimate the benefits on patients’ quality and quantity of life.

### Ethics Statement

Information about the procedure were freely available to the public domain. The research including ethics was approved by the University Research Committee (2014 n° DF84A9) and ethical approval was also obtained by the Dean of the Clinical Directorate (n°2053 20/4/2013 Chief Institutional Board of the Policlinic-Vittorio Emanuele Hospital). Written informed consent was obtained prior to study begin, and all patients signed a written consent form relating to the possible risks and benefits of the procedures. Data were acquired in compliance with GDPR regulation (General Data Protection Regulation, European Union 2016/679) and the study was retrospectively registered (ISRCTN12709516).

## Data Availability

All data produced in the present study are available upon reasonable request to the authors

## Acknowledgements

this study is dedicated to all Pioneer Sicilian Patients affected with MPM who volunteered accepted the operation of VATS PD and HITHOC at the University of Catania hospital, and to those who cannot be operated. In the memory of those who could not be helped. We thank the surgeons, doctors, nurses, radiographers, pathologists at the participating centers. We thank Policlinico “Rodolico” for financial support covering the cost of hyperthermic intrathoracic chemiotherapy.

*Collaborators PubMed citable Study Group on Malignant Pleural Mesothelioma: alphabetic order

Marco Aiello

Marinella Astuto

Massimo Caiozzo

Giovanna Fantaci

Tommaso Nicolosi

Hector Soto Parra

## Authors’ Statement

MM conceived the trial and is the chief investigator. MF, MN, RT, MF contributed to the protocol and design of the study. MC, TN, SC are sites principal investigators. TF calculated the sample size. MM wrote the original draft and MF, RT, TF, MN, SC, MF contributed to subsequent editing. MF conducted the statistical analysis. All authors reviewed and approved the final version of the paper. The authors accept full responsibility for the overall content of this manuscript. The trial database was held at Policlinico “Rodolico” Hospital.

## Technical appendix, Data Statement Section

Data can be accessed on request to

## Data Sharing

Unpublished data can be accessed on request to

## Article Summary

### Evidence before this study

Treatments options for malignant pleural mesothelioma are often palliative and only few have intention to cure. There is abundant evidence to affirm that pleuropneumonectomy is nowadays rarely performed because the reported higher mortality and because similar or even better long-term results can be obtained with less invasive surgeries such as pleurectomy/decortication (P/D) or talc pleurodesis (TP) alone. Pleurectomy/decortication is a lung sparing resection, which permits the macroscopic removal of the tumor, and has a low surgical mortality rate with a significant risk of local recurrence. Hyperthermic Intraoperative Thoracic Chemotherapy (HITHOC) is a type of adjuvant local treatment performed in the operating room immediately after surgery for MPM. Although there are many studies reporting debulking surgery plus HITHOC for the treatment of MPM, most of them all retrospective. No studies compare VATS P/D plus HITHOC with VATS talc pleurodesis for MPM.

### Added Value of this study

This study shows that the overall survival time increased in favour of VATS P/D plus HITHOC with a median survival of 28 months and with 30% of patients still alive at the time of writing. It is essential to note that in Group B, the median survival time in the 9 patients with epithelioid MPM undergoing VATS P/D plus HITHOC, was 45 months, versus 15 months in the 9 patients with epithelioid MPM undergoing VATS talc pleurodesis. It is certainly not new that patients with epithelioid MPM survive longer than those with other types of MPM, but the long-term results from our study are encouraging for the future treatment of this aggressive cancer.

### Implications of the available evidence

This pilot study represents an important step forward demonstrating the feasibility for a multicenter randomized trial to compare VATS P/D plus HITHOC with VATS talc pleurodesis in the management of MPM, and to estimate the benefits on patients’ quality and quantity of life.

